# Use of HFNC in COVID-19 patients in non-ICU setting: Experience from a tertiary referral centre of north India and a systematic review of literature

**DOI:** 10.1101/2021.06.23.21259045

**Authors:** Anivita Aggarwal, Umang Arora, Ankit Mittal, Arunima Aggarwal, Komal Singh, Animesh Ray, Manish Soneja, Pankaj Jorwal, Neeraj Nischal, Akhil Kant Singh, Puneet Khanna, Naveet Wig, Anjan Trikha

## Abstract

**Introduction:** The rapid surge of cases and insufficient numbers of intensive care unit (ICU) beds have forced hospitals to utilise their general wards for administration of non-invasive respiratory support including HFNC(High Flow Nasal Cannula) in severe COVID-19. However, there is a dearth of data on the success of such advanced levels of care outside the ICU setting. Therefore, we conducted an observational study at our centre, and systematically reviewed the literature, to assess the success of HFNC in managing severe COVID-19 cases outside the ICU.

**Methods:** A retrospective cohort study was conducted in a tertiary referral centre where records of all adult COVID-19 patients (≥18 years) requiring HFNC support were between September and December 2020 were analysed. HFNC support was adjusted to target SpO2 ≥90% and respiratory rate ≤30 per min. The clinical, demographic, laboratory, and treatment details of these patients were retrieved from the medical records and entered in pre-designed proforma. Outcome parameters included duration of oxygen during hospital stay, duration of HFNC therapy, length of hospital stay and death or discharge. HFNC success was denoted when a patient did not require escalation of therapy to NIV or invasive mechanical ventilation, or shifting to the ICU, and was eventually discharged from the hospital without oxygen therapy; otherwise, the outcome was denoted as HFNC failure. Systematic review was also performed on the available literature on the experience with HFNC in COVID-19 patients outside of ICU settings using the MEDLINE, Web of Science and Embase databases. Statistical analyses were performed with the use of STATA software, version 12, OpenMeta[Analyst], and visualization of the risk of bias plot using robvis.

**Results:** Thirty-one patients receiving HFNC in the ward setting, had a median age of 62 (50 – 69) years including 24 (77%) males. Twenty-one (68%) patients successfully tolerated HFNC and were subsequently discharged from the wards, while 10 (32%) patients had to be shifted to ICU for non-invasive or invasive ventilation, implying HFNC failure. Patients with HFNC failure had higher median D-dimer values at baseline (2.2 mcg/ml vs 0.6 mcg/ml, p=0.001) and lower initial SpO2 on room air at admission (70% vs 80%, p=0.026) as compared to those in whom HFNC was successful .A cut-off value of 1.7 mg/L carried a high specificity (90.5%) and moderate sensitivity (80%) for the occurrence of HFNC failure. Radiographic severity scoring as per the BRIXIA score was comparable in both the groups(11 vs 10.5 out of 18, p=0.78). After screening 98 articles, total of seven studies were included for synthesis in the systematic review with a total of 820 patients, with mean age of the studies ranging from 44 to 83 years and including 62% males. After excluding 2 studies from the analysis, the pooled rates of HFNC failure were 36.3% (95% CI 31.1% – 41.5%) with no significant heterogeneity (*I*^2^ =0%, p=0.55).

**Conclusions:** Our study demonstrated successful outcomes with use of HFNC in an outside of ICU setting among two-thirds of patients with severe COVID-19 pneumonia. Lower room air SpO2 and higher D-dimer levels at presentation were associated with failure of HFNC therapy leading to ICU transfer for endotracheal intubation or death. Also, the results from the systematic review demonstrated similar rates of successful outcomes concluding that HFNC is a viable option with failure rates similar to those of ICU settings in such patients.

## Introduction

The outbreak of viral pneumonia caused by Severe Acute Respiratory Syndrome Coronavirus 2 (SARS-CoV-2) originating in China in late December 2019 was declared a global pandemic by the World Health Organisation (WHO) in March 2020. After one year in December 2020, WHO had reported that the cumulative number of cases had exceeded 65 million with over 1.5 million deaths.[1] As of May 2021, India bears the world’s second largest case-burden having encountered over 20 million confirmed cases and over 322,000 deaths, with a further alarming rise in the numbers as per a WHO situation report.[2] While majority of the cases are asymptomatic or have a mild disease, around 14-20% patient develop moderate to severe disease, and roughly 5% cases become critically ill. The mortality rates among the critically ill have been found to be as high as 50-80% despite the highest level of care.[3] Given the spate of COVID-19 cases faced by nations such as India, the total number of critically ill cases, especially during the peaks, has overwhelmed its healthcare infrastructure. Maintaining oxygenation and adequate respiratory support with the help of non-invasive devices like nasal cannula, face mask, non-rebreathing mask, high flow nasal cannula (HFNC) and non-invasive ventilation (NIV) have been the pillars of management of this deadly disease. [4] The rapid surge of cases and insufficient numbers of intensive care unit (ICU) beds have forced hospitals to utilise their general wards for administration of non-invasive respiratory support including HFNC in severe COVID-19. However, there is a dearth of data on the success of such advanced levels of care outside the ICU setting. Therefore, we conducted an observational study at our centre, systematically reviewed the literature, to assess the success of HFNC in managing severe COVID-19 cases outside the ICU.

## Methodology

This was a retrospective cohort study conducted at our tertiary referral centre located in north India. Between September and December 2020, during a period of increased case burden, a large number of COVID-19 patients with severe hypoxemia were managed in our medical ward. Case records of all adult patients (≥18 years) who required HFNC support were reviewed after obtaining institutional ethical approval. Those who received HFNC after weaning from invasive mechanical ventilation or NIV, required to be transferred to the ICU within 24 hours of admission, or with incomplete medical records related to the outcome parameters were excluded from the study.

Patients with severe COVID-19 pneumonia admitted to the ward were initiated respiratory support via HFNC, if after a trial of high-flow oxygen (face mask up to 10L/min followed by non-rebreathing mask [NRBM] up to 15 L/min), they failed to achieve SpO2 of ≥90% or a respiratory rate (RR) of ≤30 breaths per min (bpm). HFNC was initiated with adequate humidification, temperature between 31° -37°C, flow rates between 30 to 60 L/min, and fraction of inspired oxygen (FiO2) of up to 100% adjusted to target SpO2 ≥90% and respiratory rate ≤30 per min. If the patient’s vital parameters worsened including inability to achieve desired SpO2 or RR, altered sensorium, development of hypercapnia or hemodynamic instability; patients were shifted to the ICU for initiation of NIV or invasive mechanical ventilation as per the discretion of the treating physician and were denoted as HFNC failure. All patients received anticoagulation (prophylactic dose enoxaparin, dalteparin, or unfractionated heparin) and steroids as per the national treatment guidelines for management of severe COVID-19 disease.

The clinical, demographic, laboratory, and treatment details of these patients were retrieved from the electronic and physical medical records, reviewed from their hospital admission till death or discharge and entered in pre-designed proforma. Demographic profile, their comorbidities, lowest room air SpO2 at admission, initial SpO2 /FiO2 ratio (S/F ratio) at the initiation of HFNC, laboratory parameters at admission etc were noted. The severity of pneumonia on chest radiograph at the time of admission were assessed by a qualified radiologist according to the *BRIXIA* score, an 18 point semi-quantitative assessment of lung disease in patients with COVID-19, that ranks the pulmonary involvement based on the extent and characteristics of radiographic abnormalities.[5] Outcome parameters included duration of oxygen during hospital stay, duration of HFNC therapy, length of hospital stay and death or discharge. HFNC success was denoted when a patient did not require escalation of therapy to NIV or invasive mechanical ventilation, or shifting to the ICU, and was eventually discharged from the hospital without oxygen therapy; otherwise, the outcome was denoted as HFNC failure.

We also systematically reviewed the available literature on the experience with HFNC in COVID-19 patients outside of ICU settings using the MEDLINE, Web of Science and Embase databases. All articles published in the English language, up till 24th December 2020, were searched using the following exploded Medical Search Headings terms and test words ((‘’COVID-19’’) OR (‘’COV-2’’)) OR (‘’Coronavirus-2’’)) OR (‘’coronavirus’’)) AND ((((‘’High Flow Nasal Cannula’’)) OR (‘’High Flow Nasal Oxygenation’’)) OR (‘’HFNC’’)) OR (‘’HFNO’’)) AND ((((‘’outside ICU’’)) OR (‘’Non-ICU’’)) OR (‘’ward’’)). All study designs (case reports and series, retrospective and prospective observational studies, and trials) found in the search, and the bibliography of these studies after manual review, were reviewed independently for eligibility by two reviewers (AA and AR). The studies were included in the review if they fulfilled the following criteria: adult (age ≥ 18 years) COVID-19 patients managed in wards or similar non-ICU settings with high flow nasal cannula therapy at any time during their hospital course. Editorials, reviews, and studies in a language other than English were excluded. Data was extracted independently by the reviewers (AA and AR)and entered in a spreadsheet. The following study characteristics were noted: study population characteristics, and place of HFNC initiation, markers of disease severity including predictors of HFNC failure (including mean SpO2/FiO2 ratio), and outcome parameters (duration of HFNC therapy, HFNC failure, escalation of respiratory support to NIV or invasive ventilation, ICU transfer or death, and hospital length of stay). In case of disagreements in study selection between the two reviewers, a third reviewer was consulted (AM). Although the included studies depicted cohorts of patients, they lacked control arm without exposure to HFNC, and thus we utilized the Joanna Briggs Institutes’ (JBI) inventory for case series to assess risk of bias in these studies.[6]

Continuous variables were summarized as means and standard deviations or medians and interquartile ranges, depending on the normality of distribution. Categorical variables were represented by frequencies and percentages. Analysis was done using a two-sample *t*-test for parametric variables, Wilcoxon rank-sum test for non-parametric variables, and Fisher exact test for categorical variables. A two-tailed P value of less than 0.05 was considered to indicate statistical significance. For meta-analysis, probability of HFNC success were pooled using a binary random effects model for proportions. Statistical analyses were performed with the use of STATA software, version 12, OpenMeta[Analyst], and visualization of the risk of bias plot using robvis as depicted in figure 3 and table 4.[7][8]

## Results

Thirty-one patients received HFNC in the ward setting during the study period, with a median (IQR) age of 62 (50 – 69) years including 24 (77%) males. Diabetes mellitus (n=16, 51%) and hypertension (n=15, 48%) were the most frequent comorbidities (Table 1). The median initial SpO2 was 75% (IQR, 67 – 84%) at room air upon presentation to the hospital emergency. All patients (n=31) were managed with HFNC and the median SpO2/FiO2 ratio was 192 (IQR, 172 – 217) at the time of HFNC initiation.

**Table 1.** Patient details

| Patient characteristics | Total | HFNC success n=21(%) | HFNC failure n=10(%) | p value |
| --- | --- | --- | --- | --- |
| Age (years), median (IQR) | 62 (50 – 69) | 60 (50 – 68) | 64.5 (53 – 72) | 0.41 |
| Male , n(%) | 24 (77.4) | 16 (76) | 8(80) | 0.81 |
| Comorbidities , n(%) |  |  |  |  |
| No comorbidities | 10 (32%) | 9 (42%) | 1 (10%) | 0.1 |
| Hypertension | 15(48.4) | 9 (42.3) | 6 (60) |  |
| Diabetes | 16(51.6) | 10(47.6) | 6 (60) |  |
| Coronary artery disease | 6(19.3) | 3 (14.3) | 3 (30) |  |
| Chronic lung disease | 2(6.6) | 1 (4.7) | 1 (10) |  |
| Malignancy | 5(16.1) | 3 (14.3) | 2 (20) |  |
| Post-transplant | 1(3) | 1 (4.7) | 0 (0) |  |
| Chronic kidney disease | 1(3) | 1 (4.7) | 2 (20) |  |
| Initial SpO2 (%), median (IQR) | 75 (67 – 84) | 80 (70 – 84) | 70 (65 – 74) | 0.036 |
| SpO2/FiO2 ratio (%), median (IQR) | 192 (172 – 217) | 196 (188 – 217) | 182 (170 – 211) | 0.25 |
| Chest radiograph severity score, median (IQR) | 9 (5 – 12) | 11 (8 – 15) | 10.5 (8 – 16) | 0.78 |
| Initial inflammatory markers |  |  |  |  |
| CRP (mg/dL), median (IQR) | 10.37 (1.76 – 13.85) | 10.81 (2.56 – 13.85) | 8.96 (0.74 – 12) | 0.52 |
| Ferritin (ng/mL), median (IQR) | 552.8 (338.1 – 1056.4) | 605.4 (339.6 – 1061.3) | 502.95 (219.4 – 844.3) | 0.29 |
| IL-6 (IU/mL), median (IQR) | 30.3 (14.23 – 87.59) | 33.69 (14.41 – 103.6) | 22.31 (14.18 – 49.96) | 0.31 |
| D dimer (mg/L), median (IQR) | 0.8 (0.41 – 2.13) | 0.6 (0.4 – 0.9) | 2.175 (1.7 – 3) | 0.0014 |

Twenty-one (68%) patients successfully tolerated HFNC, were weaned off oxygen support and subsequently discharged from the wards. They required HFNC for a median of 9 (IQR, 5 – 12) days, and oxygen therapy was required for a median of 14 days (IQR, 11 –22) days during admission. However, 10 (32%) patients had worsening respiratory parameters and had to be shifted to ICU for non-invasive or invasive ventilation, implying HFNC failure; details of which are given in table 2. Patients with HFNC failure had higher median D-dimer values at baseline (2.2 mcg/ml vs 0.6 mcg/ml, p=0.001) and lower initial SpO2 on room air at admission (70% vs 80%, p=0.026) as compared to those in whom HFNC was successful. D-dimer at presentation predicted HFNC failure well with area under ROC curve of 0.86 (95% CI 0.69-1.0). A cut-off value of 1.7 mg/L carried a high specificity (90.5%) and moderate sensitivity (80%) for the occurrence of HFNC failure, leading to correct classification in 87% of cases. The difference in other inflammatory markers was not found to be significant (table 1). Radiographic severity scoring as per the BRIXIA score was comparable in both the groups(11 vs 10.5 out of 18, p=0.78).[5] Among those who failed the HFNC trial, 9 (90%) patients required invasive mechanical ventilation in the ICU and subsequently died, while a single patient was managed with NIV in the ICU and eventually discharged (Table 2).

**Table 2.** Management and outcomes of the patients

| Management and outcome | Total | HFNC success (n=21) | HFNC failure (n=10) | p value |
| --- | --- | --- | --- | --- |
| Remdesivir use , n(%) | 16 (51.6%) | 12 (57.1%) | 4(40%) | 0.44 |
| Methylprednisolone dose per day (mg) , median (IQR) | 80 (80 – 120) | 81 (80 – 80) | 82 (80 – 120) | 0.38 |
| HFNC duration , median days (IQR) | 9 (5 – 12) | 8 (5 – 11) | 10 (6 – 17) | 0.25 |
| Required mechanical ventilation , n(%) |  | - | 9(90%) |  |
| Duration of mechanical ventilation , n(%) |  | - | 5 (3 – 9) |  |
| Total days on oxygen therapy, median days (IQR) | 14 (11 – 22) | 14 (11 – 21) | 20.5 (14 – 23) | 0.08 |
| Duration of hospital stay, median days (IQR) | 17 (14 – 25) | 16 (14 – 25) | 19.5 (14 – 26) | 0.56 |
| Mortality , n(%) | 9 (29%) | 0 | 9 (90%) | <0.001 |
| Discharged , n(%) | 22 (71%) | 21 (100%) | 1 (10%) |  |

Search through the three databases described above yielded 98 articles, which were screened and subsequently 91 studies were excluded. A total of seven studies were included for synthesis in the systematic review (Figure 1) with a total of 820 patients, with mean age of the studies ranging from 44 to 83 years, and including 62% males.[9][10][11][12][13][14] Studies were one each from the following nations Netherlands, Italy, China, France, South Africa and two from USA with the mean risk of HFNC failure ranging from 25–75%, and mortality among the studies ranging from 15% to 55%. The initial SpO2/FiO2 ratios, ROX index, duration of HFNC use, and length of hospital stay are shown in table 3. We identified failure of HFNC (defined as a composite of those who required invasive or non-invasive mechanical ventilation by NIV, transfer to the ICU, or died) from the included studies. The pooled probability of HFNC failure was 46.7% (95% CI 42.7% – 50.7%) although the results were significantly heterogeneous (*I*^2^ =88.7%, p<0.001).

**Figure 1.**
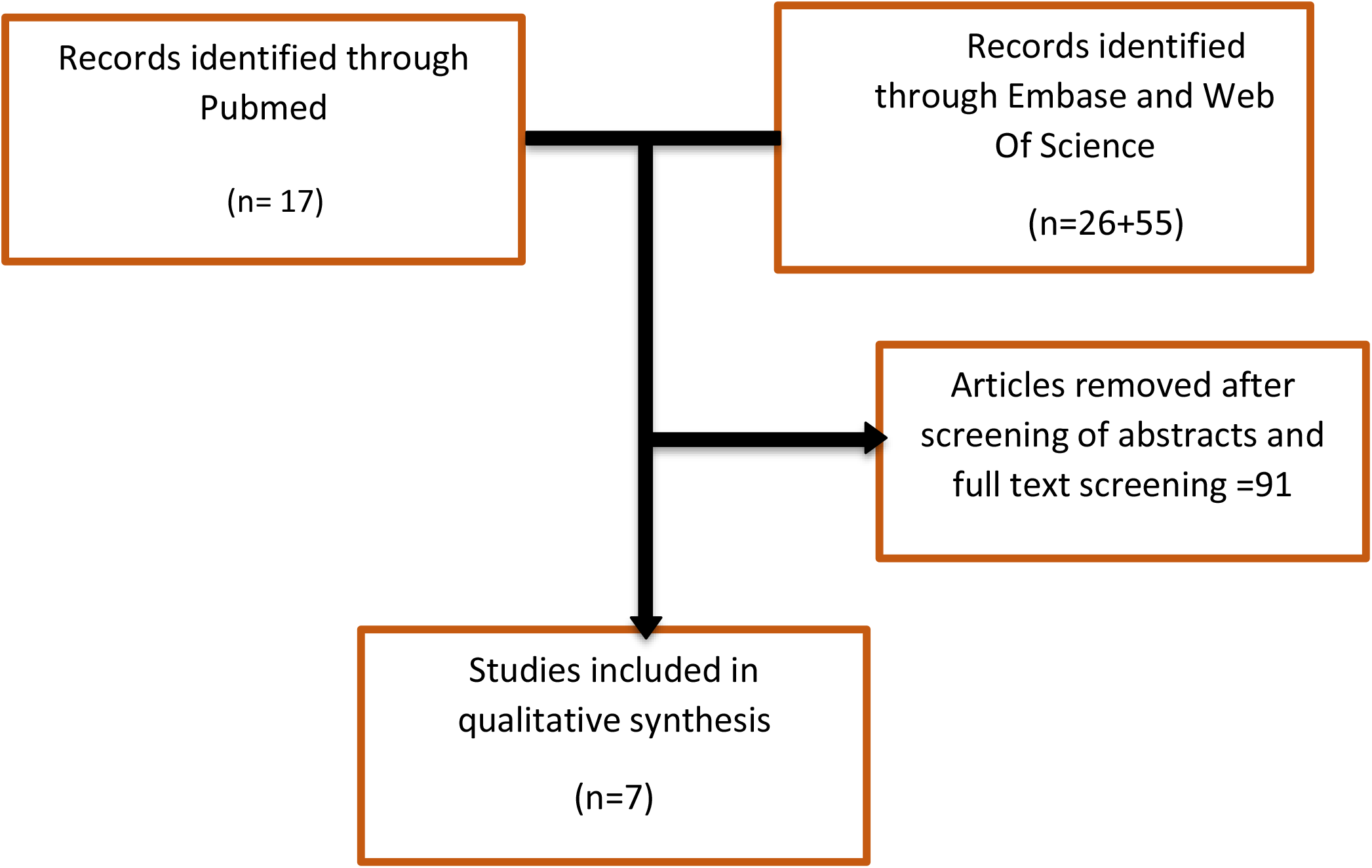
Study selection flowchart

**Table 3:** summary table of the systematically reviewed studies

| No. | First author<br>Country<br>Study design | Study population (N, Age, Sex) | Severity of hypoxemia | Duration of HFNC use | Hospital length of stay (LOS) | Need for invasive mechanical ventilation (IMV) or mortality |
| --- | --- | --- | --- | --- | --- | --- |
| 1 | Steenkiste *et al* [14]<br>Nov 2020<br>Netherlands<br>Retrospective cohort<br>(Pre-print was identified at the time of literature review, while final publication was available at the time of manuscript writing) | N=32<br>Median age 79 (74-83) years<br>Males=22(69%)<br>Frail patients with DNI instructions despite respiratory failure | Median SpO<sub>2</sub>/FiO<sub>2</sub> ratio 157.5 (150-163) | Median 9.5 days (4.8-17.5) in survivors and 2.0 days (1.0-3.0) in the non-survivors | Median LOS 15 days in survivors and 4.9 days in non-survivors | None were intubated (DNI order)<br>Mortality in 24 (75%) |
| 2 | Chandel A *et al* [13]<br>December 2020<br>USA<br>Retrospective observational | N=272<br>Mean age 57(± 13) years<br>Males= 180 (66%) | Mean SpO<sub>2</sub> 93%<br>ROX index at 2,6 and 12 hours = (4.5,4.6 and 4.7) | Median duration 3 days (1-6 days) | Total hospital stays for all not given<br>Length of ICU stay – mean 14(8,21)days | IMV in 108(39.7%) patients; 61 of these within 48 hours<br>Mortality in 49(45%)<br>60.3%(164) had success<br>Early HFNC failure – 56%(61), late HFNC failure in 47(43%) |
| 3 | Franco *et al*[12]<br>July 2020<br>Italy<br>Pre-print | N= 163 (24.3% of 690 patients were on HFNC)<br>Mean age 60(±13) years<br>Males=114(69.9%) | Mean PaO<sub>2</sub>/FiO<sub>2</sub> 166(±65) | Not mentioned | Length of hospital stay 19.2 (±13.3) days | DNI in 12/163<br>IMV in 47 (28.8%)<br>Mortality in 26 (16.9%)<br>HFNC failure in total 89 (55%) patients |
| 4 | Ming Hu *et al* [10]<br>Dec 2020<br>China | N=105<br>Mean age 65 years<br>Males= 51 (48%) | PaO<sub>2</sub>/FiO<sub>2</sub> 116 at start of HFNC<br>SpO<sub>2</sub>/FiO<sub>2</sub> and ROX (done at 2,6,12,24 hours comparing failure and success ) | Median duration 5.0 days (2.5–9.0 days) | Hospital stay – 14 days(10.5–19 days) | Mortality = 16/105<br>HFNC failure in 40 (38.1%)<br>Young age, female sex, and lower SOFA score were associated with HFNC success<br>ROX index at 6 hours had good predictive capacity for HFNC outcomes (AUROC, 0.798) |
| 5 | Guy *et al* [11]<br>July 2020<br>France | N=27 total<br>Median age 77(77-79)<br>Males= 22/27(81%) | Median PaO<sub>2</sub>/FiO<sub>2</sub> ratio at presentation 203, and at the time of HFNC initiation 124 | Median duration 6 days (2- 10 days) | Median 17 days (14-22 days) | Mortality = 4 (15%)<br>HFNC failure in 7 (25%)<br>19 patients weaned from HFNC |
| 6 | Jackson *et al* [22]<br>October 2020<br>USA<br>Abstract only | N=41 (of 70 total patients on HFNC, remaining 29 were admitted to ICU)<br>Population characteristics of all 70 patients: Mean age 64 (±18 years)<br>Males= 45(64%) U | - | - | - | Separate information for ward and ICU patient outcomes were not available. In Among all 70 patients: Mortality in 24(34%)<br>Transferred from ward to ICU 18(26%)<br>Failure of HFNC in 21/70 |
| 7 | Calligaro *et al*[9]<br>South Africa<br>October 2020 | N=188 (of total 293 patients, remaining 105 were admitted to ICU)<br>Population characteristics of all 293 patients: Median age 52(44-58) years<br>Males 163 (56%) | Median PaO<sub>2</sub>/FiO<sub>2</sub> was 68 (54-92)<br>Lower in those who had HFNC failure (63 [51–83] vs 76 [58–102], p <0.001) | Median duration 6 days (3–9 days) in HFNC success and 2 days(1–5 days) in HFNC failure | - | HFNC failure in 112/188 (59.6%) in ward and 156/293 (53%) overall<br>Mortality = 130 ( of 271 patients who had died or discharged [48%] while 22 patients had not attained at outcome)<br>Survival to hospital discharge was 128/129 (99%) and 11/140 (7%) in those with HFNC success and failure, respectively |

DNI= ‘Do not intubate’ order

**Table 4:**
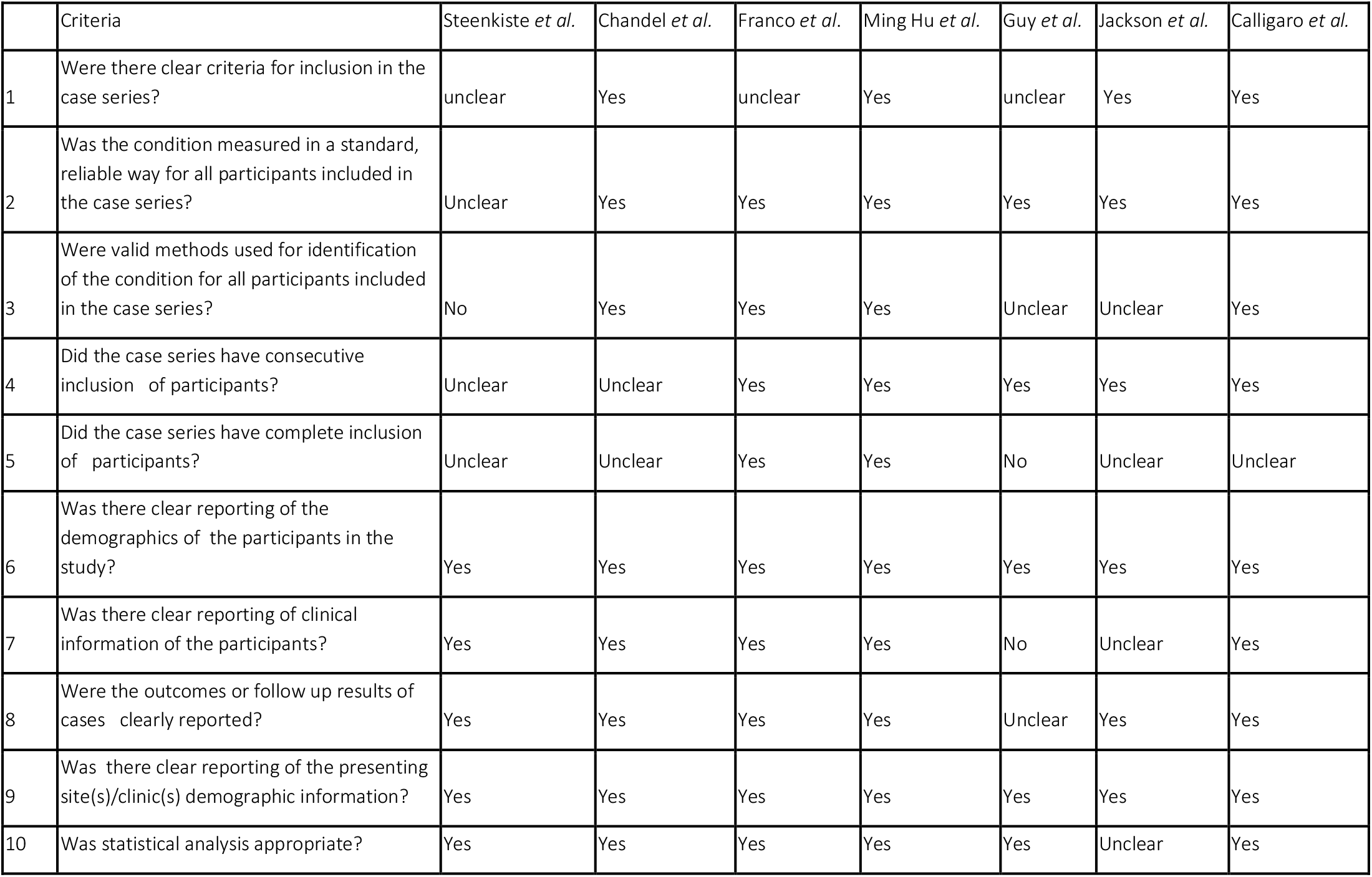
Risk of bias assessment of the studies in systematic review

|  | Criteria | Steenkiste *et al.* | Chandel *et al.* | Franco *et al.* | Ming Hu *et al.* | Guy *et al.* | Jackson *et al.* | Calligaro *et al.* |
| --- | --- | --- | --- | --- | --- | --- | --- | --- |
| 1 | Were there clear criteria for inclusion in the case series? | unclear | Yes | unclear | Yes | unclear | Yes | Yes |
| 2 | Was the condition measured in a standard, reliable way for all participants included in the case series? | Unclear | Yes | Yes | Yes | Yes | Yes | Yes |
| 3 | Were valid methods used for identification of the condition for all participants included in the case series? | No | Yes | Yes | Yes | Unclear | Unclear | Yes |
| 4 | Did the case series have consecutive inclusion of participants? | Unclear | Unclear | Yes | Yes | Yes | Yes | Yes |
| 5 | Did the case series have complete inclusion of participants? | Unclear | Unclear | Yes | Yes | No | Unclear | Unclear |
| 6 | Was there clear reporting of the demographics of the participants in the study? | Yes | Yes | Yes | Yes | Yes | Yes | Yes |
| 7 | Was there clear reporting of clinical information of the participants? | Yes | Yes | Yes | Yes | No | Unclear | Yes |
| 8 | Were the outcomes or follow up results of cases clearly reported? | Yes | Yes | Yes | Yes | Unclear | Yes | Yes |
| 9 | Was there clear reporting of the presenting site(s)/clinic(s) demographic information? | Yes | Yes | Yes | Yes | Yes | Yes | Yes |
| 10 | Was statistical analysis appropriate? | Yes | Yes | Yes | Yes | Yes | Unclear | Yes |

The heterogenity could not be explained by age or sex distribution alone. Two studies reported high rates of HFNC failure. Steenkiste et al recruited older patients (median age 79 years) patients with a do not intubate (DNI) order and had provided HFNC to patients with an indication for endotracheal intubation in accordance with patient and family wishes[14]; second, the study population of Calligaro et al included the most severely hypoxemic patients (median PaO2/FiO2 ratio 68 mmHg) belonging to severe ARDS, which conventionally require endotracheal intubation and mechanical ventilation, muscle paralysis, and prone position ventilation.[9] After excluding these 2 studies from the analysis, we found pooled rates of HFNC failure of 36.3% (95% CI 31.1% – 41.5%) with no significant heterogeneity (*I*^2^ =0%, p=0.55) as shown in figure 2.

**Figure 2:**
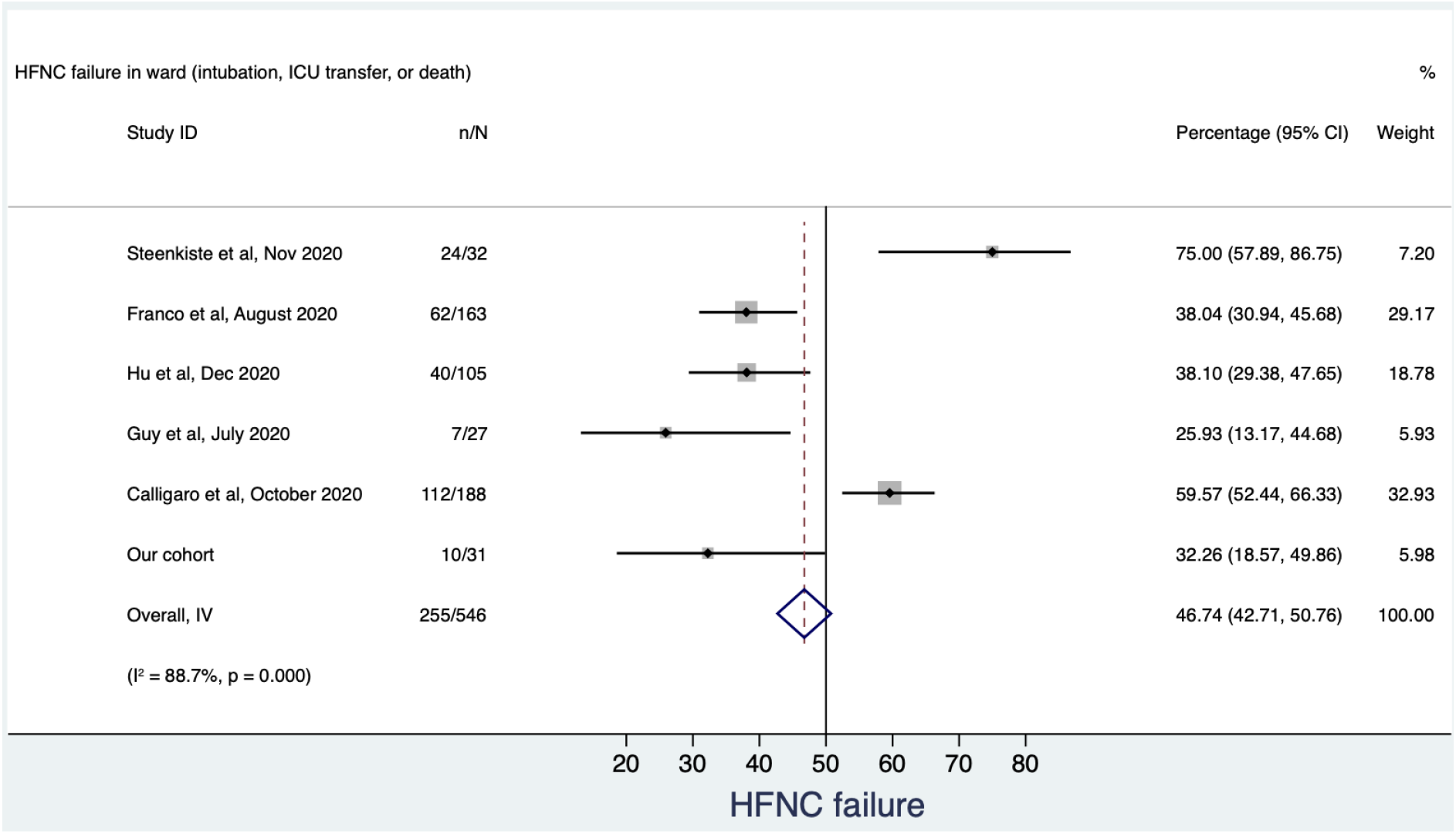
Pooled prevalence of HFNC failure defined as a composite of need for invasive mechanical ventilation and death from studies identified in the systematic review and our cohort.

**Figure 3:**
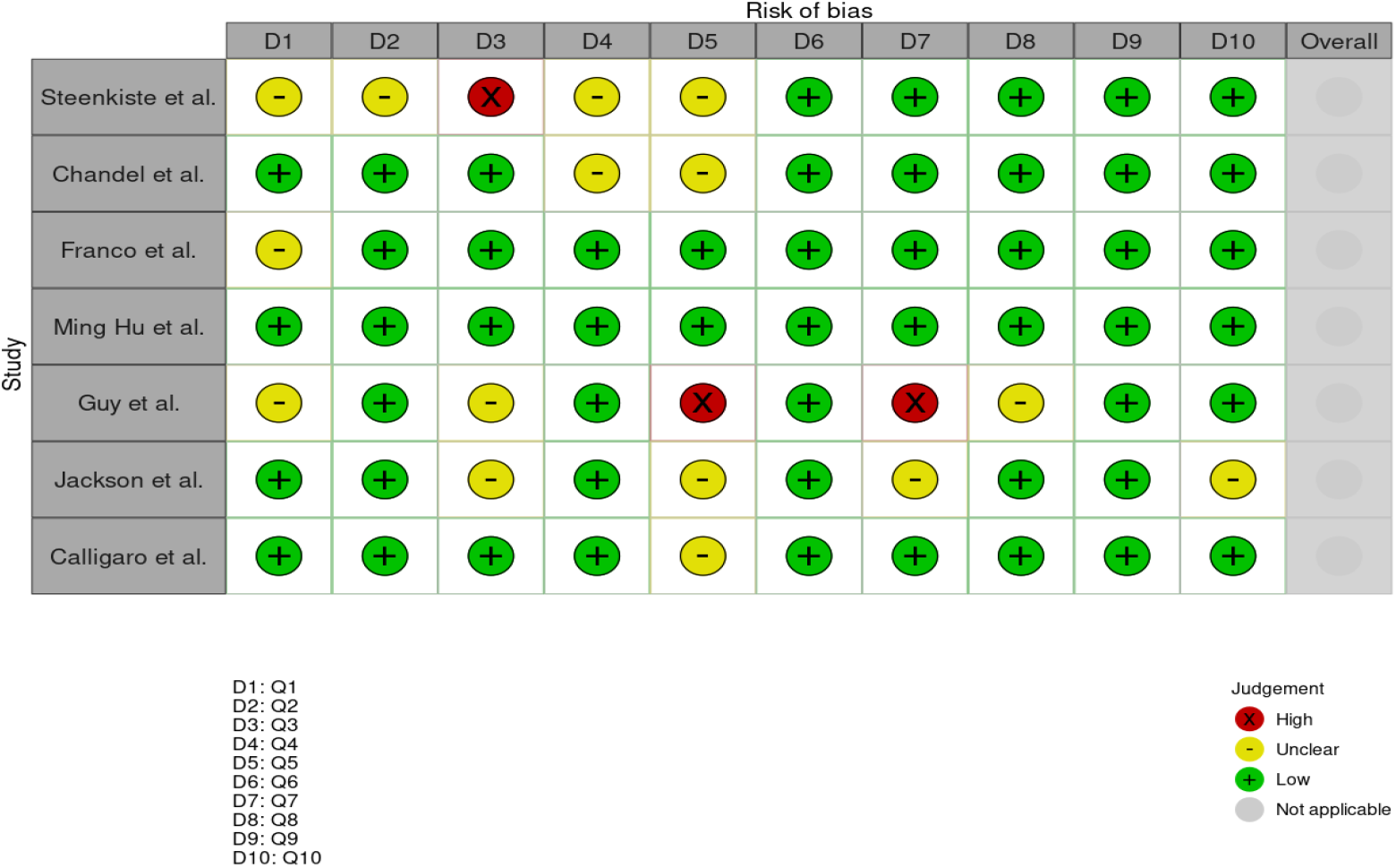
Risk of bias analysis via ROBVIS scale tool of the studies in systematic review

## Discussion

In hospitals with high influx of COVID-19 admissions and overburdened critical care units, HFNC use in wards could be a silver lining for the patients suffering from severe respiratory compromise awaiting ICU care. Our retrospective cohort demonstrated successful outcomes with use of HFNC in an outside of ICU setting among two-thirds of patients with severe COVID-19 pneumonia. Lower room air SpO2 at presentation and higher D-dimer levels at presentation were associated with failure of HFNC therapy leading to ICU transfer for endotracheal intubation or death.

Ours is the first study from India, and second from Asia, describing the outcomes of HFNC use in a non-ICU setting among COVID-19 patients. Our patient belonged to the category of severe COVID-19 with moderate to severe ARDS, and 67% availability (n=21) had a successful outcome with HFNC usage and eventual discharge to home care. Despite limited ICU beds, two-thirds patients who could not be managed with maximal NRBM oxygen support, were successfully managed using HFNC in a referral centre burdened with a heavy load of severe COVID-19 patients.

A meta-analysis studying acute hypoxemic respiratory failure has found that HFNC, NIV mask or helmet are efficacious in reducing need for endotracheal intubation compared with standard oxygen therapy.[15] They further evaluated the devices and found that helmet NIV was superior to both face mask NIV as well as HFNC in reducing mortality as well as preventing need for intubation, however both face mask NIV and HFNC carried similar rates of mortality as well as intubation. Helmet NIV is sparsely available in resource-poor settings and often needs more intensive monitoring. HFNC devices are comfortable, do not interfere with meals and speech, or cause pressure ulcers like face mask NIVs. On the other hand, they are unable to measure tidal volumes, deliver higher levels of PEEP or pressure support during inspiration. All major society guidelines deliberating on the use of oxygenation and ventilation devices in COVID-19 have advised use of HFNC in preference to NIV masks.[16] However, failure of either device is an indication for endotracheal intubation to avoid excessive delay in intubation.

Identification of individuals in whom respiratory failure can be managed with HFNC in the ward with low-risk for requirement of NIV or invasive mechanical ventilation would be instrumental for resource-limited hospitals for triaging patients to the ICU. In our study, low SpO2 at presentation as well as elevated D-Dimer were associated with HFNC failure. Also, inflammatory markers including CRP and D-dimer have been found to be more frequently elevated in severe COVID pneumonia, correlating inversely with respiratory function at presentation, and predicting mortality in COVID ARDS.[17] Similarly, retrospective cohorts studying patients in ICU[18] from Japan have found higher levels of D-dimer in those with HFNC failure compared with HFNC success [4.8 mg/L vs. 2.6 mg/L, p = 0.02], as well as ward settings such as Calligaro et al (1.03 mg/L vs 0.56 mg/L, p =0.002).[9] We found a D-dimer level of ≥1.7 mg/L to correctly classify 87% of HFNC failure cases and thus may help to triage patients at high likelihood of HFNC failure to early ICU transfer and a lower threshold for endotracheal intubation. Interestingly, D-dimer value of 2 mg/L was found to be an optimal cut-off of judging death in a Chinese cohort of COVID-19 patients from Wuhan. [19] Our present study reinforces the importance of D-dimer in predicting poor prognosis in COVID-19 patients, albeit in the backdrop of HFNC failure.

Predictably, patients with severe COVID-19 pneumonia who have lower oxygen saturation, denoting more extensive pulmonary parenchymal or pulmonary vasculature involvement, are at higher risk of mortality.[20] Consistent with this, in our study, we found median SpO2 at presentation in the emergency department to be higher in those with HFNC success (80% vs 70%, p=0.036). Several other tools have been used to predict failure of HFNC particularly the ROX index which has shown promise as its role has been replicated in several studies in diverse settings.[21] However the most appropriate cut-off value for ROX to optimally predict HFNC failure varies widely in in ward (ROX cut-off of 2.2[9], 5.5 [22]and 5.9[23]) as well as ICU settings (4.8[24] and 5.37[25]). Unfortunately, the retrospective nature of the present study did not allow us to track the ROX consistently in all our patients.

Limited sample size and a retrospective study design are major limiting factors of our study for determining the early predictors of HFNC failure conclusively. Our institute, being a tertiary referral care hospital of north India, caters to patients with severe disease, complications, and more comorbidities, providing a potential selection bias. The monitoring, doctor-patient and nurse-patient ratio, facilities in wards are much inferior to that of an ICU care setting but they may be variable in various other hospitals of resource limited countries and hence the outcomes and results may not be generalisable to other settings. Lack of access to frequent arterial blood gas analysis and CT scans made the severity assessment and patient monitoring inferior as compared to various other published studies.

In the systematic review of the 7 available observational studies, we found an overall pooled HFNC failure rate of 46.7% in hypoxemic respiratory failure due to COVID-19. However, on analysis of the heterogeneity, after excluding the two studies (described in results) we found a homogenous population of middle age, male predominant adults with moderate ARDS (according to PaO2/FiO2, or SpO2/FiO2 as a surrogate) with a pooled failure rate of 36.3%. Thus, the review authors conclude that HFNC is a viable option with failure rates similar to those of ICU settings in such patients. The review was limited by the retrospective nature of the included studies as well as the small number of patients in the majority of them. The included patients among the studies differed with regards to the age, severity of hypoxemia at presentation, medical treatment protocol, intubation thresholds, as well ethical constraints (such as DNI order).A multicentric retrospective cohort reviewed 324 patients from ICUs in Wuhan, China and found relatively high HFNC failure rates of 55%, which was associated with age > 60 years, ROX index <5.31, thrombocytopenia, and elevated IL-6 values. [26] Calligaro et al compared outcomes of HFNC in wards vis-a-vis ICU and found that the HFNC failure rate experienced in ICU (44/105, 41.9%) was lower than in wards (76/188, 59.6%).[9] In another study from an Intermediate Respiratory Centre in Madrid (Spain) using HFNC in COVID-19 associated ARDS, a failure rate of 52.5% was noted with overall mortality rate of 22.5% .[27] The higher rate of HFNC failure in ward settings reiterates that HFNC use, like all high flow oxygen therapy, has better outcomes in an intensive setup. Their conclusion that patients could be managed effectively with HFNC in the ward was borne out of necessity due to a major uptick in COVID-19 cases they experienced, similar to what India has faced in April – May 2021.

## Conclusions

High flow nasal cannula has widely been used in the COVID-19 hypoxemic respiratory failure. In this study we found HFNC use outside ICU settings to be feasible, with a failure rate of approximately 32% in patients with severe COVID-19 pneumonia which is similar to the failure rate found in the systematic review (36%). HFNC failure in our cohort was predicted well by D-dimer at presentation, with a cut-off of 1.7 mg/L having a positive predictive value of 80%. HFNC use is ideally to be applied in ICU settings, however in these unprecedented times of overwhelming influx of patients in hospitals, HFNC use in wards could be a glimmer of hope for the patients awaiting ICU admissions.

## Data Availability

The study data can be accessible on request via email on the corresponding author email address

---

**Supplementary Figure 1:**
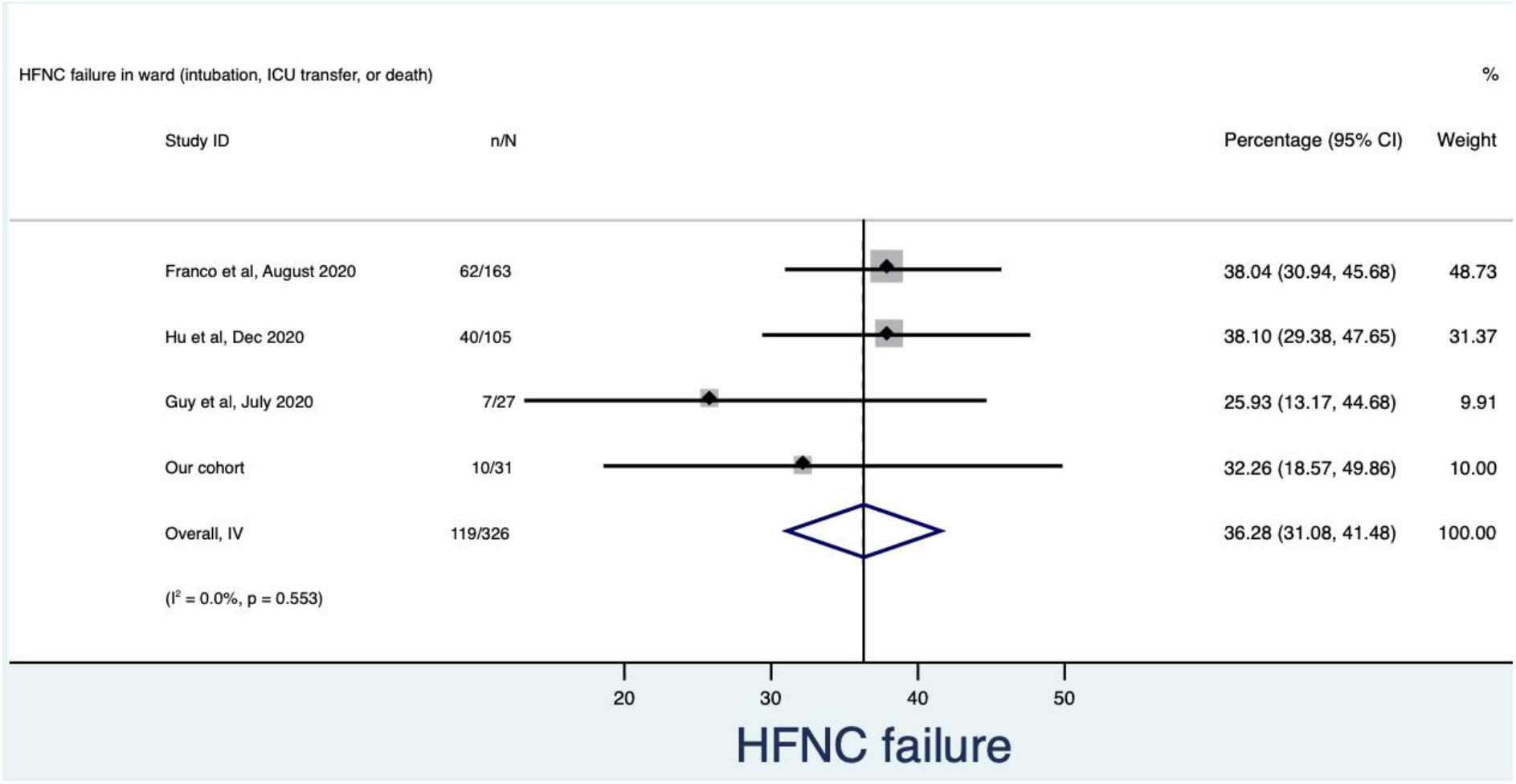
Pooled prevalence (%) of HFNC failure defined as a composite of need for invasive mechanical ventilation and death from studies identified in the systematic review and our cohort, after excluding two studies (for details see *Results*)

## Notes

### Competing Interest Statement

The authors have declared no competing interest.

### Funding Statement

None

### Author Declarations

Institute ethics committee, All India, Institute of Medical Sciences, New Delhi, India 110029 provided ethical approval as "Approved from ethical angle prospectively w.e.f. 20th Jan 2021" under reference number IEC-51/08.01.2021, RP-11/2021. Decision made:ethical approval given

